# Decision trees for COVID-19 prognosis learned from patient data: Desaturating the ER with Artificial Intelligence

**DOI:** 10.1101/2022.05.09.22274832

**Authors:** Nikolas Bernaola, Guillermo de Lima, Miguel Riaño, Lucia Llanos, Sarah Heili-Frades, Olga Sanchez, Antonio Lara, Guillermo Plaza, Cesar Carballo, Paloma Gallego, Pedro Larrañaga, Concha Bielza

## Abstract

**Objectives:** To present a model that enhances the accuracy of clinicians when presented with a possibly critical Covid-19 patient.

**Methods:** A retrospective study was performed with information of 5,745 SARS-CoV2 infected patients admitted to the Emergency room of 4 public Hospitals in Madrid belonging to Quirón Salud Health Group (QS) from March 2020 to February 2021. Demographics, clinical variables on admission, laboratory markers and therapeutic interventions were extracted from Electronic Clinical Records. Traits related to mortality were found through difference in means testing and through feature selection by learning multiple classification trees with random initialization and selecting the ones that were used the most. We validated the model through cross-validation and tested generalization with an external dataset from 4 hospitals belonging to Sanitas Hospitals Health Group. The usefulness of two different models in real cases was tested by measuring the effect of exposure to the model decision on the accuracy of medical professionals.

**Results:** Of the 5,745 admitted patients, 1,173 died. Of the 110 variables in the dataset, 34 were found to be related with our definition of criticality (death in <72 hours) or all-cause mortality. The models had an accuracy of 85% and a sensitivity of 50% averaged through 5-fold cross validation. Similar results were found when validating with data from the 4 hospitals from Sanitas. The models were found to have 11% better accuracy than doctors at classifying critical cases and improved accuracy of doctors by 12% for non-critical patients, reducing the cost of mistakes made by 17%.

## Introduction

The effects of the Covid-19 pandemic have overwhelmed the resources of the public medical system around the world, with saturation of medical resources being one of the most concerning side-effects. As long as herd immunity is not reached through mass vaccination, the risk of a wave of infection (Fig. 1) that swamps hospitals again and stretches resources even thinner would be disastrous.

**Fig. 1:**
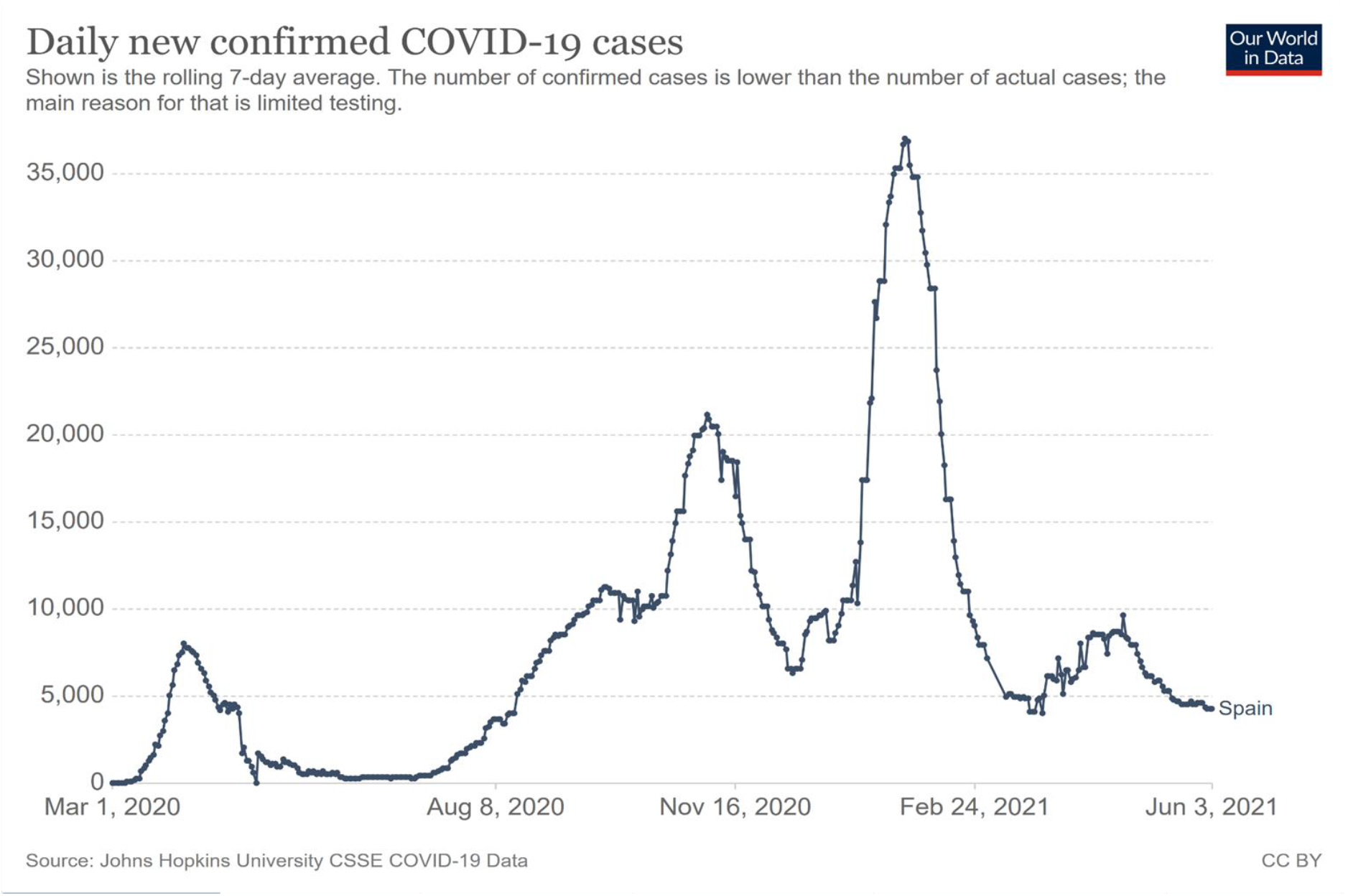
Daily new confirmed cases of COVID-19 in Spain. (Data by Johns Hopkins University, chart by Our World in Data (Roser, 2021))

Several models have been developed using different approaches to predict the evolution of Covid-19 infection including a study with Spanish patients (Berenguer et al., 2021) predicting 30-day mortality and building a checklist for ease of use or low-resource approaches using patients’ histories instead of data during admission (Estiri et al., 2021). However, various literature reviews (Wynants et al., 2020; Roberts et al., 2021) show that most models are at a risk of bias and overfitting, since they do not generalize well to data from other hospitals (because of small datasets or poor model selection), and they are not focused on being deployed in real world situations, with the review by Roberts and colleagues finding that none of the models reviewed were fit for clinical use. These problems make most models never achieve their stated goal of helping diagnose or predict patient evolution. Taking these problems to heart, we built a decision tree classifier with data from 5,745 patients from 4 public Hospitals in Madrid belonging to the Quirón Salud Health Group. Our work tackles the problems mentioned by the reviews head on by prioritizing explainability, with clear indicators of why the tree is making every decision so that the medical professional can choose to use it or ignore it with full information. Furthermore, since the cost of a false negative is much higher than a false positive, we have not optimized purely for accuracy but have made a choice to penalize false negatives higher. Finally, thinking about deployment of the model, we have validated the performance of the model not only on its own through cross-validation and a test set from four different hospitals belonging to another organization (Sanitas Hospitals Health Group) but also relative to medical professionals with the same information and in combination with them to see if our method helps them make better decisions.

The results of this work are two models to predict Covid-19 infection severity. The first one uses only the data at admission (patient demographic data, vitals, and previous conditions) while the second one adds data from laboratory tests.

## Methods

### Source of data

Permission was obtained from local Ethics Committee (CEIm-FJD) to perform an observational, retrospective study. De-identified data were extracted from electronic Clinical Records in 5,745 SARS-CoV2 infected patients attending 4 public Hospitals in Madrid belonging to the Quirón Salud Health Group from the beginning of the pandemic (24/02/2020) to 23/02/2021. The dataset consists of 277,332 entries, with analytic data from different days from each of 7,351 unique patients from four hospitals from the FJD group.

A validation dataset was obtained through the Sanitas Hospital network which contributed data from 975 patients and four hospitals admitted from 24/02/2020 to 15/11/2020. This dataset was used to test generalization of the model.

### Filtering the data

The QS dataset contains every patient that attended one of the four hospitals during the time period and was diagnosed with Sars-Cov-2 after a positive PCR. For our analysis we decided to keep only patients who had been admitted to the hospitals, reducing the total number to 5,745. Then, for these patients, we only took the entries for which they had at least one measured analytical variable, giving us 45,625 in total.

### Missing data

All variables except for the initial laboratory tests and initial vital signs were complete in the original dataset. The ones that were not and were over the 50% completion threshold were imputed using an iterative imputation procedure from scikit-learn in Python (Buck, 1960; Pedregosa et al., 2011).

### Problem statement

#### Outcome

The first question the doctors in the team were interested in answering was whether a patient was in a critical state and needed urgent care. We chose a threshold of 72 hours and labelled the patients as critical if the time from when the sample was taken to death was less than the threshold. To build the dataset used for training the model, we used, for each of the patients that died, the first critical entry in chronological order. For the patients that did not die, we chose the first sample after admission. This led to a final dataset of 5,745 samples of which 1,173 died and 4,572 did not.

#### Predictors

Variables were chosen from the full dataset to build two models. For the first one, we only used data that would usually be available at admission in the ER (Table 1a) and which have already been associated with death in Spanish Covid-19 patients (Berenguer et al., 2020) and in our dataset for a total of 13 variables. For the second model, we added data from laboratory tests and reduced the 110 laboratory variables in the dataset to 24 through feature selection by learning 1000 trees with random initialization of the samples and selecting only the variables that were used at least once by the decision trees to separate between critical and non-critical patients (Table 1b). (Appendix A shows the full list of variables with their mean and interquartile range)

**Table 1.** Predictor variables used for the ER model and the laboratory test model. Table 1a, on the left, shows the ER variables. These were found to be associated with all-cause mortality through pairwise hypothesis tests. Table 1b, on the right, shows the extra laboratory variables added to the second model and the fraction of times they appeared in our feature selection procedure.

| EC Model Variables |  |
| --- | --- |
| **Demographics** | Age |
|  | Sex |
| **Initial vitals** | O2 Saturation |
|  | Body temperature |
|  | BMI |
| **Comorbidities** | Smokers |
|  | Cardiovascular |
|  | Pulmonary |
|  | Diabetes |
|  | Renal |
|  | Neurologic |
|  | Oncologic |
|  | Hypertension |

**Table 1. Predictor variables used for the ER model and the laboratory test model.**
| Lab test model variables |  |
| --- | --- |
| Variable name | Importance |
| RDW | 1 |
| Albumin | 1 |
| RBC count | 1 |
| Hemoglobin | 1 |
| Lymphocyte % | 1 |
| Neutrophil count | 1 |
| Segmented neutrophils % | 1 |
| Glomerular filtration rate (MDRD4) | 1 |
| Urea | 1 |
| Total protein | 0.99 |
| Lactate | 0.96 |
| CHCM | 0.95 |
| Leukocyte count | 0.93 |
| PCO2, gas | 0.86 |
| Hematocrit | 0.81 |
| Sodium | 0.69 |
| Chlorine | 0.62 |
| Fibrinogen | 0.55 |
| LDH | 0.16 |
| Creatinine | 0.09 |
| D-Dimer | 0.09 |
| Ph, gas | 0.08 |
| Interleukin 6 | 0.03 |
| Monocyte % | 0.01 |

#### Importance of the problem

The doctors chose to try to answer the question of criticality as a proxy for whether a given patient should be admitted to the hospital when the number of available beds is low. In ideal circumstances where admission does not have a cost, every patient for which there is reasonable doubt of the prognosis or who might benefit from stay would be admitted. When the hospital is saturated due to a rise in cases and due to resource constraints, some patients can follow treatment at home.

This is why a decision system that can be adjusted depending on the relative cost of admission or rejection of critical and non-critical patients and that leads to increased accuracy in diagnosis would be very useful to better allocate resources and reduce the workload, especially during the more demanding times when the hospital is saturated.

The decision to make two separate models was based on the actual procedure of deciding whether a patient should or should not be admitted. First, the doctor encounters the new patient and has access to limited information through exploration and the history of the patient. They must decide based on this limited information, and that is what the ER model is trying to support. If this decision is not made with enough confidence, they usually ask for more tests, including a laboratory test. Once the results arrive, they update their original decision with the new information, and that is what the laboratory test model is imitating. By separating the decision into two steps, we reduce the necessity of asking for extra information when it might not increase the confidence of the decision and so reduce the stress on limited laboratory capacity and streamlining decision making under pressure.

## Methods

### Decision trees

A decision tree is a simple machine learning model that consists of a series of nodes and edges, starting from a single root to multiple leaves (Quinlan, 1986). At each node, the decision tree has a sample of the patients and every sample at the node is classified as the majority class with probability estimated by the relative frequency of the class. Then, at each node except the leaves, the classifier chooses a variable and a cut-off point: samples below the cut-off will be sent to the left child of the node and samples over the cut-off will be sent to the right. The process continues until there is a minimum number of samples in a node.

Each new sample then gets classified according to the set of rules the tree describes. Starting from the root, we apply each of the cut-off values to the variables of interest and go down a path until we reach a leaf at which point the sample is classified according to the majority class.

We learned these trees through an evolutionary algorithm (using the *evtree* package in R. Grubinger et al., 2012) where we chose parameters that made the loss due to a false negative (sending a critical patient home) higher than that of a false positive (admitting a non-critical patient to the hospital). The weight between these two types of errors can be changed to learn a new model that considers the current situation at the hospital. We used relative weights of 3:1 (false negative vs false positive) as a first approximation after consideration with the medical team and carried out a sensitivity analysis on different weights and their effects on model metrics. (See validation in the next section).

### Advantages of decision trees

Our main concern when choosing a model for this task was making it as transparent as possible for the experts. Since it was going to aid and supplement decision making and never replace it we needed the reasoning of why the model was giving a choice to be as clear as possible for the doctors. This is in line with the recently released European framework proposal for regulating AI (Artificial Intelligent) (European commission, 2020), in which AI systems that deal with critical decisions in which human lives might be involved are required to explain their decision making in a way an expert can understand.

Decision trees have an inherent advantage in this respect since the decision algorithm can be interpreted directly as a set of rules. Furthermore, the output probability has a clear meaning. As an example, when a patient is said to be critical with 65% probability, that means that 65% of patients with similar characteristics were found to be critical (Fig. 2 shows an example of a branch of the laboratory test model).

**Fig. 2:**
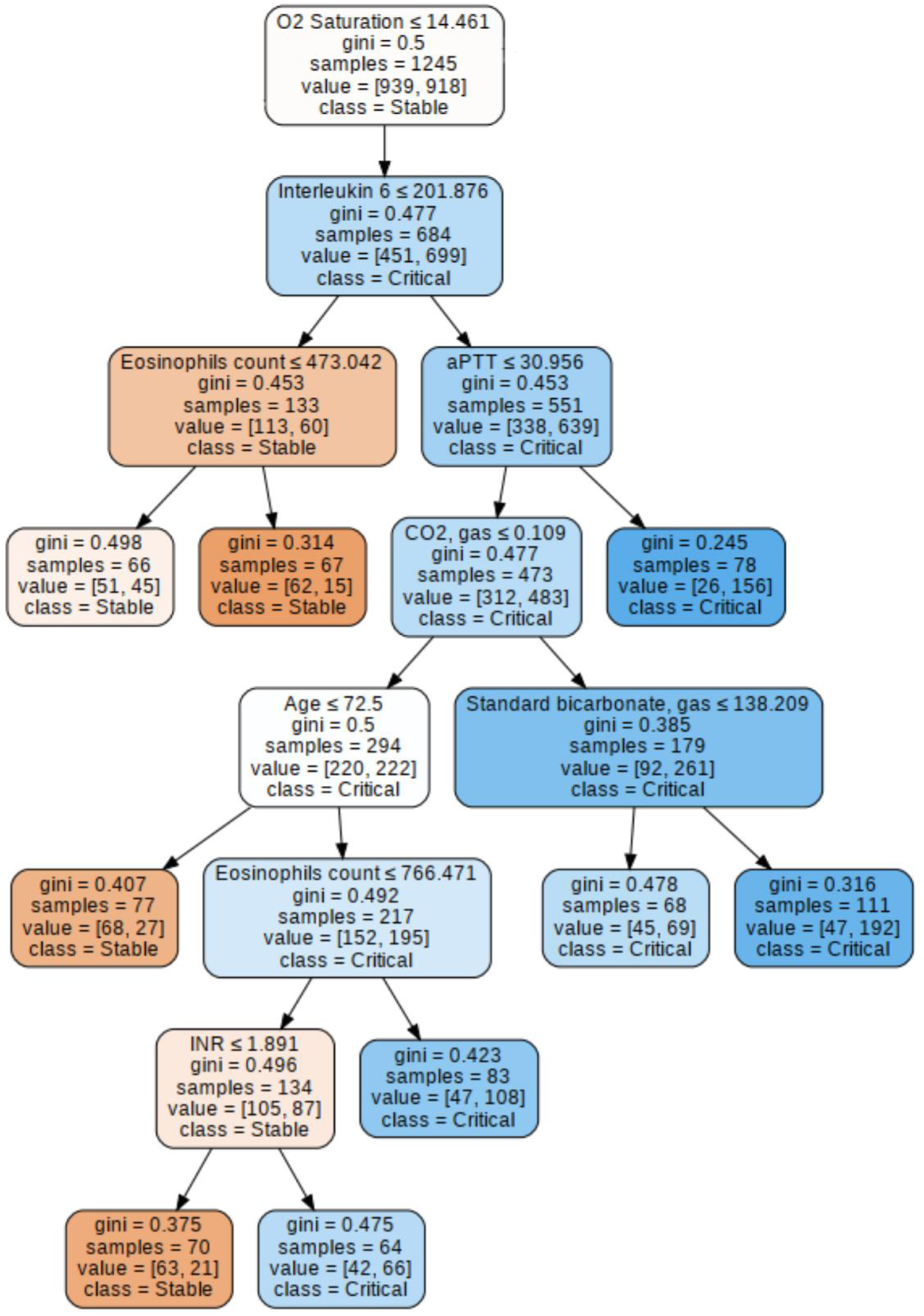
A branch of the laboratory test model. Every node has a number of patients (the samples) that are divided between stable and critical (in square brackets, first number is stable second is critical). At the top of the node there is a variable and a cut-off point. Each node sends samples that are over the cut-off to the right and the samples below to the left. The variable and cut-off are selected to try to make the children nodes as pure as possible (only having samples from one class). The purity of each node is represented visually by the color (blue for critical and orange for stable). If we have a new patient, we can classify it by following the tree depending on the actual values of each of the variables until we reach a leaf, which won’t have any children. At that point the patient is placed in the class with a majority in the leaf and the probability assigned to it is the relative frequency of the majority class in the leaf.

When AI is assisting in decision making, the relevant metric is not the accuracy of the model but by how much it improves the unaided accuracy of the decision maker (weighted by the costs of each mistake). This is why we consider that a model that explains its reasoning and can lead the decision maker to consider some parameters they might be missing is better than a black box (Price, 2018) which might make errors silently.

## Results and validation

There were two steps to the validation of this model. First, we checked that the model was fitting the data we had well and calculated the AUROC as is usual for this work, while also carrying out an analysis on the effect of different relative costs of false positives to false negatives. However, as we have mentioned before we believe that the real test of the model is if it can improve the accuracy of decision makers. To test this, we created a validation set and asked doctors to rate each patient as critical or not before and after seeing the model output, trying as much as possible to mimic the conditions under which the model would be deployed following a similar method to (Tschandl, 2020).

### Internal validation

After learning both models, we got accuracies of 83% and 85% and AUROCs were 0.76 and 0.90 for the ER and laboratory models respectively with 10-fold cross-validation. For the analytics model, which would be the last one the doctors consulted, we carried out a sensitivity analysis by relearning the model with different values of the relative costs of false positives and negatives. AUROC stayed mostly constant at 0.90 over all different weight ratios and the results for specificity, sensitivity and accuracy can be seen in figure 2. For the full table of results, see appendix B.

Almost constant AUROC shows that changing the penalties for mistakes trades off between false positive rate and false negative rate while maintaining good model performance. This can be seen in the chart above, with sensitivity decreasing as the cost of a false negative decreases while accuracy and precision increase (due to a reduced number of false positives).

### Validation from external data

Finally, with a dataset of 975 patients admitted to four hospitals from the Sanitas Hospital Network we validated generalization of the model. These patients were admitted with Covid-19 between 24/02/2020 and 15/11/2020 and were processed in the same way as the QS dataset. Figure 3 shows the results obtained by using the model learnt with the original dataset to predict the status (critical or not) of the patients in this dataset for different relative costs of mistakes (see the full data in Appendix C). The results are very similar to those of the original dataset, being even better in some cases which shows that the model can generalize to data from different centres. Using the same cost ratio we used for the original model (3:1 false negative to false positive) we get accuracies of 81.5% and 84.9% for the ER and lab model respectively (compared with 83% and 85%) and for the laboratory model we get 55% sensitivity and 49% precision compared to the 51% sensitivity and 46% precision we get with the original dataset.

**Fig. 3:**
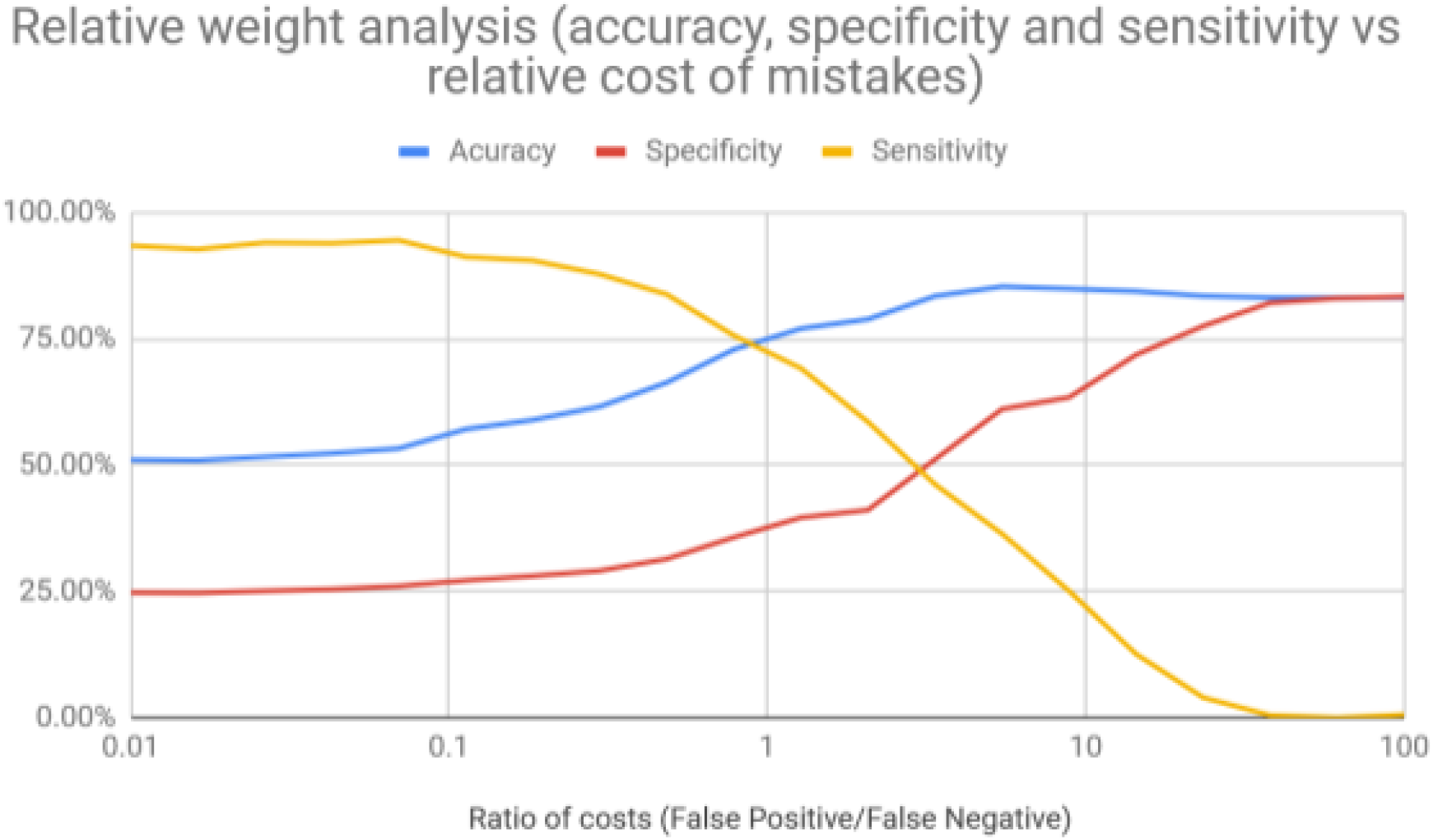
Results of the cost analysis of the laboratory test model. Shows variation of recall, precision and accuracy as the ratio of the costs of false positives and false negatives increases. Adjusting the cost and relearning the model would update its recommendations according to the current situation of the hospital.

### Validation as an aid to decision making

For the final test of the model, we randomly separated 100 patients from the QS dataset that had not been used to train the models. Anonymised data from these patients were presented to 36 doctors with different levels of experience: eleven of them were residents and twenty-five attending physicians. Presentation of the data followed two steps: first, they were given a patient knowing only its history (past conditions like cardiac conditions), age, temperature and O2 saturation. They then decided whether they think the patient is critical or not and get shown the model prediction and explanation. With the new information, they can decide to maintain their choice or change it. In the second stage they had a similar presentation but adding a table with the laboratory tests for the patient highlighted in red any variables that are outside the normality range (Fig. 5). Again, they made a choice, received the model prediction and are then able to change it. By recording the answers before and after getting the model information we can estimate the change in accuracy due to the model and see if it helps improve clinical decisions or not.

**Fig. 4:**
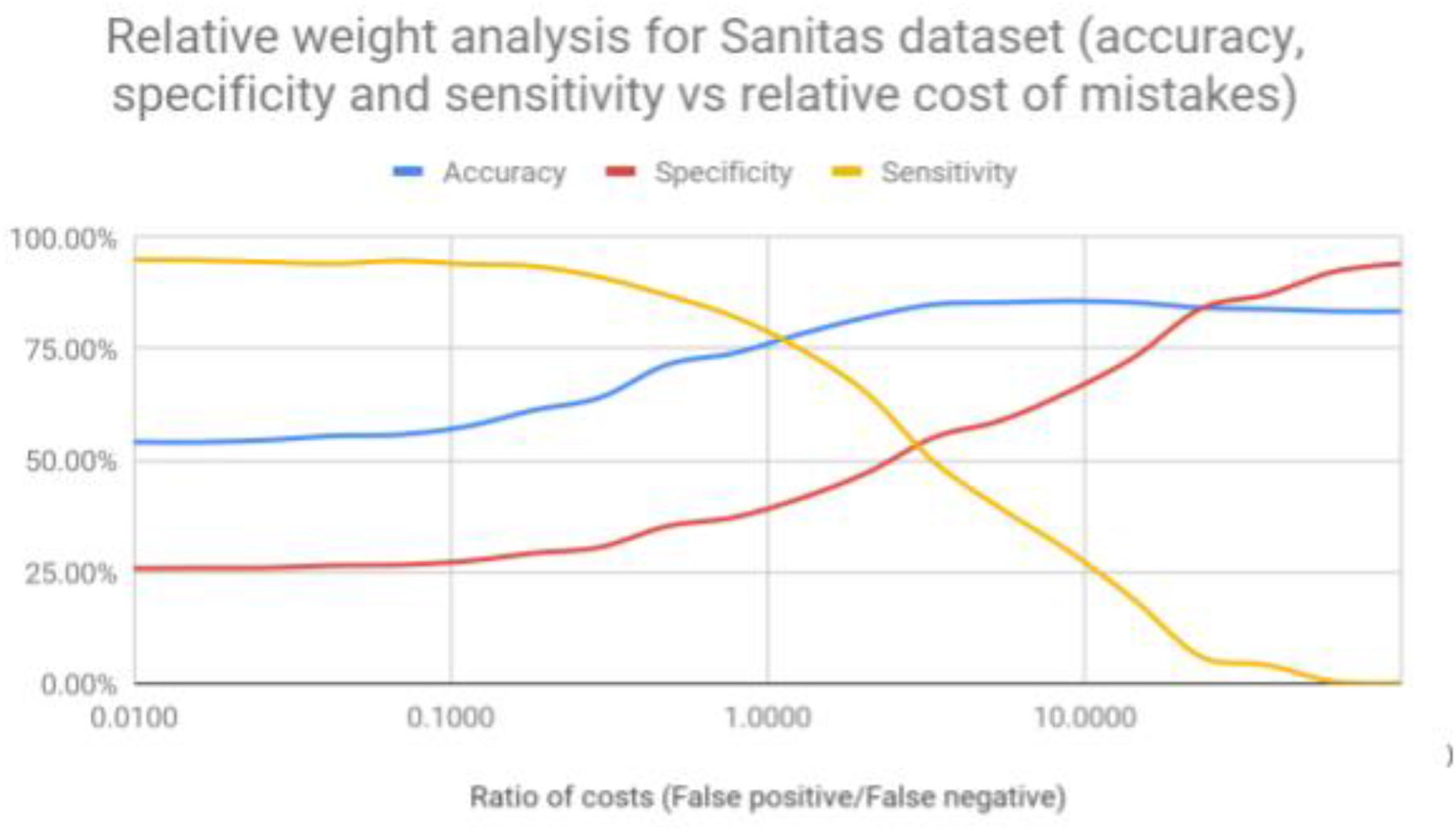
Results of the cost analysis using the laboratory test model to predict the validation dataset from Sanitas. Shows similar variation to Fig.2 and very similar results which makes us more confident on the ability of the model to generalize well to different hospitals.

**Fig. 5:**
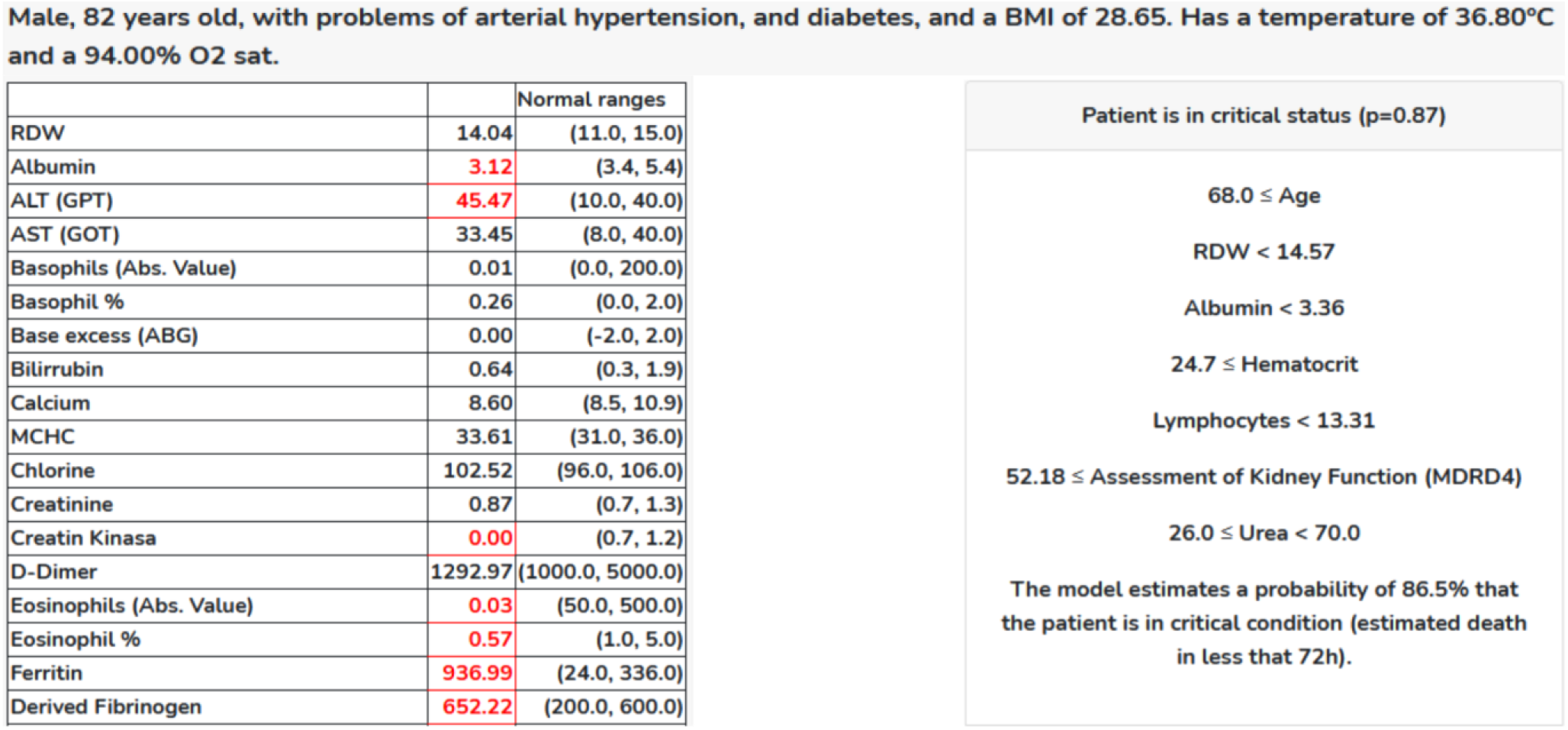
Example of model output (decission and explanation). On top, we have an example case presented in the same way it was given to doctors during validation with part of the laboratory test on the left (actual values and normality intervals given. The variables that fall outside the intervals marked in red.) On the right, we have the output of the model for this patient (critical with a probability of 86.5%) and the explanation the model gives.

The 36 doctors answered a total of 872 validation questions. The answers with and without the model were compared in three ways: comparing the accuracy, sensitivity and relative cost of mistakes and checking for significant differences between them adjusting for multiple comparisons (Dunn, 1961). For the whole validation dataset we found an improvement of 1.4% in accuracy, 0.4% in sensitivity and a reduction of costs of 2.1% all non-significant. We did subgroup analysis of attending physicians and residents and observed some differences (models seemed to help attending doctors more) but they were also not statistically significant. Finally, we analysed the patients for which the model gave a low probability of admission. These patients are the ones we are interested in for the practical application since the model has been trained to be sensitive to critical patients to avoid false negatives. Due to the extreme sensitivity, when the model is sure that the patient is stable the doctor can be very sure of the decision. Here the model performs well, with an improvement of 12% in accuracy, 1% in sensitivity and 17% in reduction of costs of mistakes all with p<0.05 after Bonferroni correction.

## Discussion

One of the most interesting aspects of the model from the clinical point of view is that it is built based on medical reasoning in its different stages. By mimicking the reasoning process of doctors, it can help them during all the steps and improve as more data is added. This was one of the priorities when designing the model, we not only needed a robust algorithm but also one that could be useful under limited data conditions. This is even more important when we consider countries where the resource scarcity is even more pronounced and where vaccination campaigns can take a much longer time.

The mortality of this pandemic, beyond the inherent to its own severity and tissue impact has been marked by the exponentiality of cases, the speed of the course of the disease and the finite capacity of hospitals. Although this capacity could be expanded at the expense of the non-Covid pathology, in many cases it was insufficient, and this was especially true in the first wave.

A model like the one we presented would have very importantly mitigated these effects in the hospital structures of all countries. But the Covid pandemic has not finished. New threats in the form of new variants lurk and although we hope that the waves will not be of the magnitude of the previous ones, a real commitment is expected in this new pandemic era in which these waves should coexist with the prepandemic activity. In this scenario it is especially important also to have a tool to classify and use the necessary resources without this existing detriment in relation to patients who will not have Covid. Additionally, the model can be adjusted to the different hospital pressure scenarios, a differential fact with respect to other models (Wynants et al., 2020).

## Limitations

The current version of the model is limited by the amount and origin of the data with which the model is trained. Both datasets, Sanitas and QS come from private hospital networks in Madrid which gives a sample that is biased towards patients that can afford private healthcare. We have been in communication with the regional government of Madrid and various public hospitals, but we have been unable to get access to the data. Furthermore, we believe that the validation results could be better with a bigger sample of doctors and with an extra group which had been trained with the tool, allowing us to observe if further training after the basic notion of how the model works would be helpful in improving the results.

## Conclusions

We show here that the development of a reliable risk-stratification tool which follows the recommendations for machine learning models (validation on outside data, easy to understand and use by the medical professionals and with transparent reasoning) in hospitals during the pandemic is feasible. The overall algorithm can be scaled to any type of unit/hospital in the world if they are collecting data. This would offer personalized results adapted to the environment of the unit analyzed. The models can be found at https://modelling-pandemics.com.

## Data Availability

All data produced in the present study are available upon reasonable request to the authors. Data used for the study is available trough contact with the corresponding hospitals due to needing ethical approval.

## Acknowledgements

This work has been partially supported by the BBVA Foundation’s grants (2020 Call) for Scientific Investigation Teams SARS-CoV-2 and COVID-19 through the “Outcome prediction and treatment efficiency in patients hospitalized with Covid-19 in Madrid: A Bayesian network approach” project, by the Spanish Ministry of Science and Innovation through the PID2019-109247GB-I00 project and through the Research Network “Artificial Intelligence in Biomedicine” (RED2018-102312-T).

The authors would also like to thank the IT teams from Sanitas and Quiron Salud as well as the medical teams without which we could not have had access to this data.

## Contributors

The following people contributed to the validation of the models:

- From the Fundacion Jimenez Diaz team: Javier Alfayete, Abulkader El Hachem, Itziar Fernandez, Iker Fernandez-navamuel, Alba Naya, Marcel Rodriguez, Maria Jesus Rodriguez, Rebeca Armenta
- From the Ramon y Cajal team: Gonzalo Aparicio, Andres Jimenez, Maria Hernandez, Ana Maria Ioan, Nuria Garcia Montes, Marta Fernandez

## Appendices

## Appendix A: Dataset description (Model variables)

| Variable name | Discharged | Deceased |
| --- | --- | --- |
| Number of patients - n (%) | 4572 (79.6) | 1173 (20.4) |
| **Age - n (%)** |  |  |
| <40 | 425 (9.3) | 4 (0.3) |
| 40-59 | 1365 (29.9) | 67 (5.7) |
| 60-79 | 1714 (37.5) | 415 (35.4) |
| >=80 | 1068 (23.4) | 687 (58.6) |
| **Female - n (%)** | 2063 (45.1) | 515 (43.9) |
| **Initial vital signs - median (IQR)** |  |  |
| Oxygen saturation - % | 94.8 (93.0-96.4) | 94.0 (91.0-96.0) |
| Body temperature - °C | 36.9 (36.5-37.2) | 36.8 (36.4-37.1) |
| BMI | 27.4 (25.0-29.3) | 26.7 (24.3-28.9) |
| **Laboratory tests - median (IQR)** |  |  |
| Lymphocyte - % | 14.3 (11.1-22.5) | 11.3 (6.7-13.7) |
| Segmented neutrophils - % | 77.6 (67.6-82.4) | 81.3 (77.0-87.9) |
| Albumin - g/dL | 3.5 (3.2-3.8) | 3.2 (2.9-3.4) |
| CHCM - g/dL | 33.7 (33.2-34.6) | 33.4 (32.6-34.0) |
| Chlorine - mmol/L | 102.0 (100.0-104.0) | 102.4 (100.0-105.6) |
| Creatinine - mg/dL | 0.8 (0.7-1.0) | 0.9 (0.7-1.3) |
| D-Dimer - ng/mL | 814.2 (381.0-1518.7) | 1519.0 (802.0-2455.0) |
| Fibrinogen - mg/dL | 577.4 (512.5-679.0) | 590.4 (519.6-703.0) |
| Glomerular filtration rate (MDRD4) - mL/min | 60.0 (59.0-60.0) | 59.0 (49.0-60.0) |
| Hematocrit - % | 37.5 (35.1-41.0) | 35.4 (33.0-40.2) |
| RBC count - 10<sup>6</sup>/μL | 4.3 (4.0-4.7) | 4.0 (3.6-4.5) |
| Hemoglobin - g/dL | 12.7 (11.7-14.0) | 11.9 (10.9-13.4) |
| Interleukin 6 | 28.9 (3.5-68.8) | 54.2 (8.0-111.8) |
| LDH - U/L | 270.2 (217.0-326.0) | 332.0 (256.0-415.0) |
| Leukocyte count - 10<sup>3</sup>/μL | 7.9 (5.5-9.5) | 8.6 (6.3-11.2) |
| Neutrophil count - 10<sup>3</sup>/μL | 5.9 (3.7-7.4) | 7.0 (4.9-9.4) |
| PCO<sub>2</sub>, gas - mmHg | 46.7 (45.4-47.9) | 47.1 (45.5-48.8) |
| Ph, gas | 7.4 (7.4-7.4) | 7.4 (7.4-7.4) |
| Total protein - g/dL | 6.2 (5.9-6.6) | 6.0 (5.6-6.3) |
| RDW - % | 13.5 (12.7-14.4) | 14.5 (13.6-15.7) |
| Sodium - mmol/L | 139.0 (137.0-141.0) | 139.0 (137.4-142.1) |
| Urea - mg/dL | 42.0 (31.0-55.0) | 61.5 (46.0-84.0) |
| Lactate - mM/L | 1.7 (1.5-2.0) | 1.9 (1.6-2.2) |
| **Comorbidities - n (%)** |  |  |
| Smokers | 264 (5.8) | 54 (4.6) |
| Cardiovascular | 967 (21.2) | 492 (41.9) |
| Pulmonary | 893 (19.5) | 271 (23.1) |
| Diabetes | 919 (20.1) | 347 (29.6) |
| Renal | 356 (7.8) | 176 (15.0) |
| Neurologic | 556 (12.2) | 251 (21.4) |
| Oncologic | 276 (6.0) | 124 (10.6) |
| Hypertension | 2104 (46.0) | 822 (70.1) |

## Appendix B: Results of sensitivity analysis

| Ratio of costs (False positive/False negative) | Accuracy | AUROC | Specificity | Sensitivity | F1-score |
| --- | --- | --- | --- | --- | --- |
| 0.01 | 51.03% | 0.901 | 24.75% | 93.40% | 0.391 |
| 0.016 | 50.91% | 0.901 | 24.70% | 92.70% | 0.390 |
| 0.026 | 51.62% | 0.903 | 25.09% | 93.99% | 0.396 |
| 0.043 | 52.32% | 0.902 | 25.41% | 93.88% | 0.400 |
| 0.070 | 53.28% | 0.903 | 26.03% | 94.46% | 0.408 |
| 0.113 | 57.11% | 0.904 | 27.18% | 91.17% | 0.419 |
| 0.183 | 58.92% | 0.906 | 28.06% | 90.46% | 0.428 |
| 0.298 | 61.57% | 0.906 | 29.10% | 87.75% | 0.437 |
| 0.483 | 66.33% | 0.904 | 31.43% | 83.75% | 0.457 |
| 0.785 | 72.87% | 0.906 | 35.71% | 75.62% | 0.485 |
| 1.27 | 77.00% | 0.905 | 39.61% | 69.14% | 0.504 |
| 2.07 | 78.85% | 0.905 | 41.11% | 58.54% | 0.483 |
| 3.36 | 83.42% | 0.900 | 51.11% | 46.17% | 0.485 |
| 5.46 | 85.31% | 0.901 | 61.07% | 36.40% | 0.456 |
| 8.86 | 84.87% | 0.901 | 63.39% | 25.09% | 0.359 |
| 14.4 | 84.37% | 0.901 | 71.81% | 12.60% | 0.214 |
| 23.4 | 83.44% | 0.906 | 77.50% | 4.00% | 0.076 |
| 37.9 | 83.14% | 0.906 | 82.16% | 0.47% | 0.009 |
| 61.6 | 83.08% | 0.906 | 83.21% | 0.00% | - |
| 100 | 83.14% | 0.904 | 83.43% | 0.59% | 0.012 |

## Appendix C: Sensitivity analysis of Sanitas hospitals

| Ratio of costs (False positive/False negative) | Accuracy | Specificity | Sensitivity | F1-score |
| --- | --- | --- | --- | --- |
| 0.0100 | 54.11% | 25.79% | 94.93% | 0.406 |
| 0.0162 | 54.07% | 25.87% | 94.83% | 0.406 |
| 0.0264 | 54.53% | 25.91% | 94.45% | 0.407 |
| 0.0428 | 55.48% | 26.42% | 94.07% | 0.413 |
| 0.0695 | 55.78% | 26.57% | 94.74% | 0.415 |
| 0.113 | 57.61% | 27.42% | 93.97% | 0.425 |
| 0.183 | 61.23% | 29.19% | 93.49% | 0.445 |
| 0.298 | 64.11% | 30.54% | 91.00% | 0.457 |
| 0.483 | 71.45% | 35.23% | 86.99% | 0.502 |
| 0.785 | 74.01% | 37.20% | 82.20% | 0.512 |
| 1.27 | 78.31% | 41.47% | 74.93% | 0.534 |
| 2.07 | 82.11% | 47.25% | 64.98% | 0.547 |
| 3.36 | 84.94% | 55.15% | 49.67% | 0.523 |
| 5.46 | 85.37% | 58.91% | 39.23% | 0.471 |
| 8.86 | 85.70% | 65.27% | 29.86% | 0.410 |
| 14.4 | 85.37% | 73.13% | 18.76% | 0.299 |
| 23.4 | 84.21% | 84.03% | 6.03% | 0.112 |
| 37.9 | 83.89% | 87.19% | 4.11% | 0.078 |
| 61.6 | 83.43% | 92.27% | 0.38% | 0.008 |
| 100 | 83.40% | 94.13% | 0.00% | - |

